# A Large Language Model Approach to Extracting Causal Evidence across Study Designs for Evidence Triangulation

**DOI:** 10.1101/2024.03.18.24304457

**Authors:** Xuanyu Shi, Wenjing Zhao, Ting Chen, Chao Yang, Jian Du

## Abstract

Health strategies increasingly emphasize both behavioral and biomedical interventions, yet the complex and often contradictory guidance on diet, behavior, and health outcomes complicates evidence-based decision-making. Evidence triangulation across diverse study designs is essential for establishing causality, but scalable, automated methods for achieving this are lacking. In this study, we assess the performance of large language models (LLMs) in extracting both ontological and methodological information from scientific literature to automate evidence triangulation. A two-step extraction approach—focusing on cause-effect concepts first, followed by relation extraction—outperformed a one-step method, particularly in identifying effect direction and statistical significance. Using salt intake and blood pressure as a case study, we calculated the Convergeny of Evidence (CoE) and Level of Evidence (LoE), finding a trending excitatory effect of salt on hypertension risk, with a moderate LoE. This approach complements traditional meta-analyses by integrating evidence across study designs, thereby facilitating more comprehensive assessments of public health recommendations.

## Introduction

It is increasingly recognized that health strategies should prioritize both behavioral interventions and biomedical interventions (e.g., medications)^1^. Social determinants of health (SDoH), especially lifestyle factors such as diet and exercise, are pivotal in managing major chronic diseases such as cardiovascular diseases, cancer, chronic respiratory diseases, and diabetes. For instances, according to data from the Institute for Health Metrics and Evaluation (IHME), behavioral factors contribute significantly to ischemic heart disease and stroke, accounting for 69.2% and 47.4% of Disability-Adjusted Life Years (DALYs), respectively—the highest among all diseases. In particular, dietary factors contributed 57.1% and 30.6% of DALYs, respectively^2^. Developing evidence-based prevention and intervention strategies encounters significant challenges due to the rapidly growing and piecemeal evidence, along with complex causal relationships from various study designs, including confounding and reverse causation. Evaluating the level of causality within a body of scientific evidence is a fundamental task, especially when research findings are inconsistent^3-5^.

Meta-analysis (META) is an effective scientific method for quantitatively synthesizing research conclusions. Utilizing statistical techniques, it combines the results of different studies to obtain an overall quantitative estimate of the impact of specific interventions (e.g., salt restriction) on particular outcomes (e.g., blood pressure). It balances conflicting evidence quantitatively to achieve evidence-based decision-making based on synthesized scientific evidence. Since its introduction in the 1970s, meta-analysis has had a significant impact on various fields such as medicine, economics, sociology, and environmental science^6^. Over the past four decades, meta-analysis has evolved to include increasingly complex methods for quantifying evidence, particularly concerning the consistency of results from the same study design or the replicability of studies. In contrast, convergency, reflecting the extent to which a given hypothesis is supported by different study designs, has not received the same attention^4^. Currently, considering consistency and convergency is recognized as an important strategy for addressing the reproducibility crisis for the scientific community^4^.

In recent years, the idea of “triangulation” has been introduced into the scientific community to measure the convergency of scientific conclusions derived from different study designs ^4, 7, 8^, particularly in human behaviours^9^. These study designs have different and independent potential sources of bias^7^. Triangulation is a research strategy involving the use of at least two research methods to investigate and analyze the same research question, mutually validating each other to enhance the robustness and reproducibility of conclusions. If conclusions derived from different research designs (such as observational studies (OS), mendelian randomization studies (MR), and randomized controlled trials (RCT), etc.) regarding the same cause-and-effect question (in fact, these study designs all aim to establish correlation) are consistent, the reliability of causality is stronger. At this point, correlation is moving towards causality. When the results point to different directions, understanding the major source of bias instruct researchers future study designs^7^.

However, current evidence triangulation studies primarily employ qualitative methods to explain the reliability of causality, lacking quantitative approaches. Researchers are accustomed to using retrospective description of relevant literature in the “Discussion” section of their papers, simply summarizing and discussing how many studies support the conclusions of the current study, how many do not, and reasons for lack of support, such as different experimental conditions^8^. A few pieces of empirical work on evidence triangulation involves a very high proportion of manual evidence screening and extraction for data elements^10^. Such retrospective, qualitative triangulation methods are susceptible to issues such as subjective selectivity of evidence and cognitive biases among different researchers.

Implementing a fully quantitative method for evidence triangulation requires a computable representation of research findings and relevant metadata obtained from different study designs. Apart from determining the presence and direction of the effects (i.e., significant increase, significant decrease, and null) between an intervention and outcome, finer-grained information of research design among many lines of evidence need to be extracted. For evidence triangulation task, it is important to extract information such as measured outcomes, effect direction of intervention (increased vs. decreased), characteristics of study populations (e.g., demographics), and other relevant contextual information.

Currently, there are natural language processing methods available for extracting conclusions from clinical research reports. This includes the utilization of Large Language Models (LLMs) to extract entities and relationships from RCT reports ^11-13^. However, these methods are predominantly based on the less specific framework of evidence-based medicine, which emphasizes Population-Intervention-Comparation-Outcome (PICO) related concepts, such as Trialstreamer and the EvidenceMap^14, 15^. While some of these methods involve effect size and direction^16, 17^, extracting and representing research design information from various sources of evidence, which is essential for triangulation, remains a subject for ongoing research. Most recently, there are attempts trying to accelerate evidence triangulation process by taking advantage of computable knowledgebase in the form of a Subject-Predicate-Object semantic triple, such as SemMedDB^18^. However, the accuracy and recall rates of medical concepts and their relationships extracted in SemMedDB are relatively low.

In this study, we try to examine the capabilities of LLMs in extracting ontological information such as intervention-outcome concepts, determining effect directions, as well as identifying methodological information such as study design. Our objective is to develop an automatic approach to aggregate various lines of SDoH-related evidence across different study designs into a computable and comparable format that is ready for quantitative evidence triangulation. We also aim to utilize the extracted data elements to assess the Convergency of Evidence (CoE, which represents the trending effect direction after triangulation) and the Level of Evidence (LoE, which denotes the strength of that direction). The overall logic and overflow of this work is shown in Figure 1.

**Figure 1.**
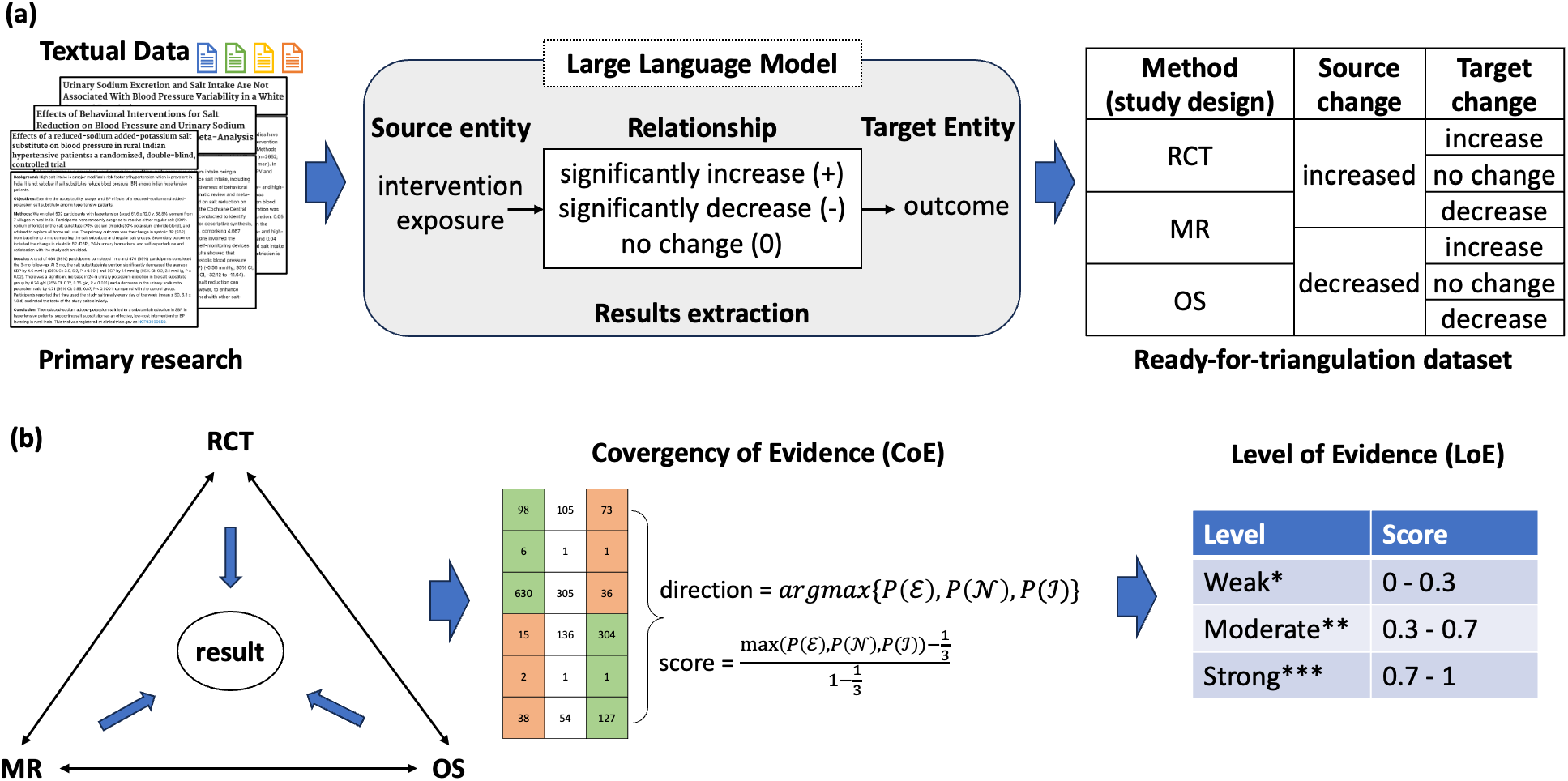
Overall workflow of automatic evidence triangulation using LLM. (a) The pipeline of using LLM to extract study designs, entities, and relationships from textual titles and abstracts. (b)The framework of evidence triangulation; The Convergency of Evidence represents the integration algorithm of the supporting and opposing evidence behind the relationship. The Level of Evidence score represents the reliability of causal relationship with scaled classification in three levels: weak (one star*), moderate (two stars**), and strong (three stars***).

## Results

### Validation of Model Performance

To validate the performance of our model in named entity recognition (NER) and relationship extraction (RE) in SDoH research, we employed a comprehensive evaluation approach using BERTScore, a state-of-the-art metric for assessing the similarity between textual representations of exposure and outcome pairs^19^. BERTScore provides precision, recall, and F1-score metrics to quantify the semantic similarity between predicted and reference text sequences during validation. Given that LLM-extracted entities may not always perfectly align with human-extracted entities, we employed this similarity-based scoring method to more rigorously assess the extent to which the LLM’s extractions correspond to the gold standard.

We compared the predicted associations generated by the model against the manually curated gold standard dataset. The following steps were undertaken:

- **Similarity Assessment:** BERTScore was calculated for each exposure and outcome pair to evaluate the semantic similarity between the model’s predictions and the gold standard. For each PMID, precision, recall, and F1-score were computed, allowing for a nuanced understanding of the model’s ability to capture relevant associations.
- **Matching and Thresholding:** We set a BERTScore threshold of 0.8 to identify matching pairs of exposures and outcomes between the predicted and gold standard data. Only those pairs exceeding this threshold were considered valid matches.
- **Evaluation of Direction and Significance:** For the matched pairs, we further evaluated the model’s performance in predicting the direction (e.g., positive, negative) and significance of the associations. Standard metrics— precision, recall, and F1-score—were calculated to quantify the model’s performance in these dimensions.
- **Error Analysis:** We identified and reported falsely predicted associations in terms of direction and significance, providing insights into areas where the model may need further refinement. This part is provided in Supplementary material #1.

### Part 1: an expert-extracted dataset of relationships between food & nutrition and cardiovascular outcomes

In the one-step one-shot extraction, GPT-4o-mini achieved the highest F1 scores for exposure (0.86) and outcome (0.82) extraction, demonstrating strong overall performance. However, glm-4-airx had slightly higher precision in extracting the direction of the relationship, although all models showed moderate performance in this category. For significance extraction, deepseek-chat and GPT-4o-mini exhibited high F1 scores (0.86 and 0.87).

The two-step extraction method generally outperformed the one-step approach, particularly in handling complex indicators like direction and significance. Deepseek-chat was the most reliable model with an F1 score of 0.82 in direction and 0.96 in significance, especially in the two-step approach.

**Table 1.** Model performance comparison on one-step and two-step extraction of entities and relationships between food&nutrition and cardiovascular outcomes.

| Model\Indicator | Exposure |  |  | Outcome |  |  | Direction |  |  | Significance |  |  |
| --- | --- | --- | --- | --- | --- | --- | --- | --- | --- | --- | --- | --- |
|  | P | R | F1 | P | R | F1 | P | R | F1 | P | R | F1 |
| **One-step extraction** |  |  |  |  |  |  |  |  |  |  |  |  |
| deepseek-chat | 0.82 | 0.83 | 0.82 | 0.82 | 0.81 | 0.81 | 0.73 | 0.71 | 0.71 | 0.80 | 0.94 | 0.86 |
| glm-4-airx | 0.86 | 0.84 | 0.85 | 0.80 | 0.79 | 0.79 | 0.73 | 0.73 | 0.73 | 0.86 | 0.81 | 0.82 |
| qwen-plus | 0.85 | 0.87 | 0.85 | 0.82 | 0.83 | 0.82 | 0.69 | 0.72 | 0.70 | 0.38 | 0.45 | 0.40 |
| GPT-4o-mini | **0.85** | **0.88** | **0.86** | **0.81** | **0.82** | **0.81** | 0.78 | 0.81 | 0.79 | 0.83 | 0.94 | 0.87 |
| **Two-step extraction** |  |  |  |  |  |  |  |  |  |  |  |  |
| deepseek-chat | 0.84 | 0.86 | 0.85 | 0.78 | 0.8 | 0.79 | **0.82** | **0.83** | **0.82** | **0.94** | **0.98** | **0.96** |
| glm-4-airx | 0.80 | 0.80 | 0.80 | 0.81 | 0.81 | 0.80 | 0.80 | 0.83 | 0.81 | 0.64 | 0.95 | 0.72 |
| qwen-plus | 0.84 | 0.87 | 0.85 | 0.79 | 0.81 | 0.80 | 0.55 | 0.55 | 0.55 | 0.82 | 0.89 | 0.84 |
| GPT-4o-mini | 0.83 | 0.87 | 0.85 | 0.78 | 0.81 | 0.79 | 0.81 | 0.82 | 0.81 | 0.95 | 0.97 | 0.96 |

### Part 2: External validation of human-extracted relationships between dietary factors and coronary heart disease

Additionally, we validated the approach in an external human-extracted dataset (validation dataset #2). In the one-step extraction approach, deepseek-chat achieved F1-scores of 0.76 for exposure, 0.88 for outcome, and 0.67 for association. Glm-4-airx performed consistently with F1-scores of 0.77 for exposure, 0.82 for outcome, and 0.79 for association. Qwen-plus showed competitive results with F1-scores of 0.76 for exposure, 0.84 for outcome, and 0.72 for association.

In the two-step extraction approach, deepseek-chat improved to F1-scores of 0.78 for exposure, 0.88 for outcome, and 0.75 for association. Glm-4-airx demonstrated balanced performance with F1-scores of 0.76 for exposure, 0.85 for outcome, and 0.86 for association. Qwen-plus maintained competitive performance with F1-scores of 0.74 for exposure, 0.88 for outcome, and 0.77 for association. Deepseek-chat and glm-4-airx emerged as the most reliable models across both extraction methods.

**Table 2.** Model performance comparison on one-step and two-step extraction of entities and relationships in the external validation dataset of effect of dietary factors on coronary heart disease.

| Model\Indicator | Exposure |  |  | Outcome |  |  | Association |  |  |
| --- | --- | --- | --- | --- | --- | --- | --- | --- | --- |
|  | P | R | F1 | P | R | F1 | P | R | F1 |
| **One-step extraction** |  |  |  |  |  |  |  |  |  |
| deepseek-chat | 0.77 | 0.77 | 0.76 | **0.89** | **0.88** | **0.88** | 0.68 | 0.71 | 0.67 |
| glm-4-airx | 0.78 | 0.76 | 0.77 | 0.80 | 0.82 | 0.80 | 0.81 | 0.79 | 0.79 |
| qwen-plus | 0.77 | 0.76 | 0.76 | 0.82 | 0.84 | 0.83 | 0.71 | 0.75 | 0.72 |
| GPT-4o-mini | 0.76 | 0.75 | 0.75 | 0.76 | 0.79 | 0.77 | 0.72 | 0.74 | 0.73 |
| **Two-step extraction** |  |  |  |  |  |  |  |  |  |
| deepseek-chat | **0.78** | **0.79** | **0.78** | 0.87 | 0.88 | 0.87 | 0.74 | 0.83 | 0.75 |
| glm-4-airx | 0.77 | 0.76 | 0.76 | 0.85 | 0.85 | 0.85 | **0.87** | **0.88** | **0.86** |
| qwen-plus | 0.76 | 0.74 | 0.74 | 0.88 | 0.88 | 0.88 | 0.75 | 0.81 | 0.77 |
| GPT-4o-mini | 0.79 | 0.79 | 0.78 | 0.81 | 0.83 | 0.82 | 0.76 | 0.82 | 0.78 |

### A case of evidence triangulation of salt on blood pressure

In our analysis, we explored the effect of salt intake on blood pressure using a dataset derived from 1,488 studies. After re-classifying the study designs using LLM model, this dataset included 476 RCTs, 5 MRs, 795 OSs and 140 Meta Analysis/Systematic Review/Review. We employed the proposed LLM pipeline to extract structured information based on the PICO framework along with primary efficacy results and other relevant metadata from these publications. A sample of the extracted evidence is presented in Figure 2.

**Figure 2.**
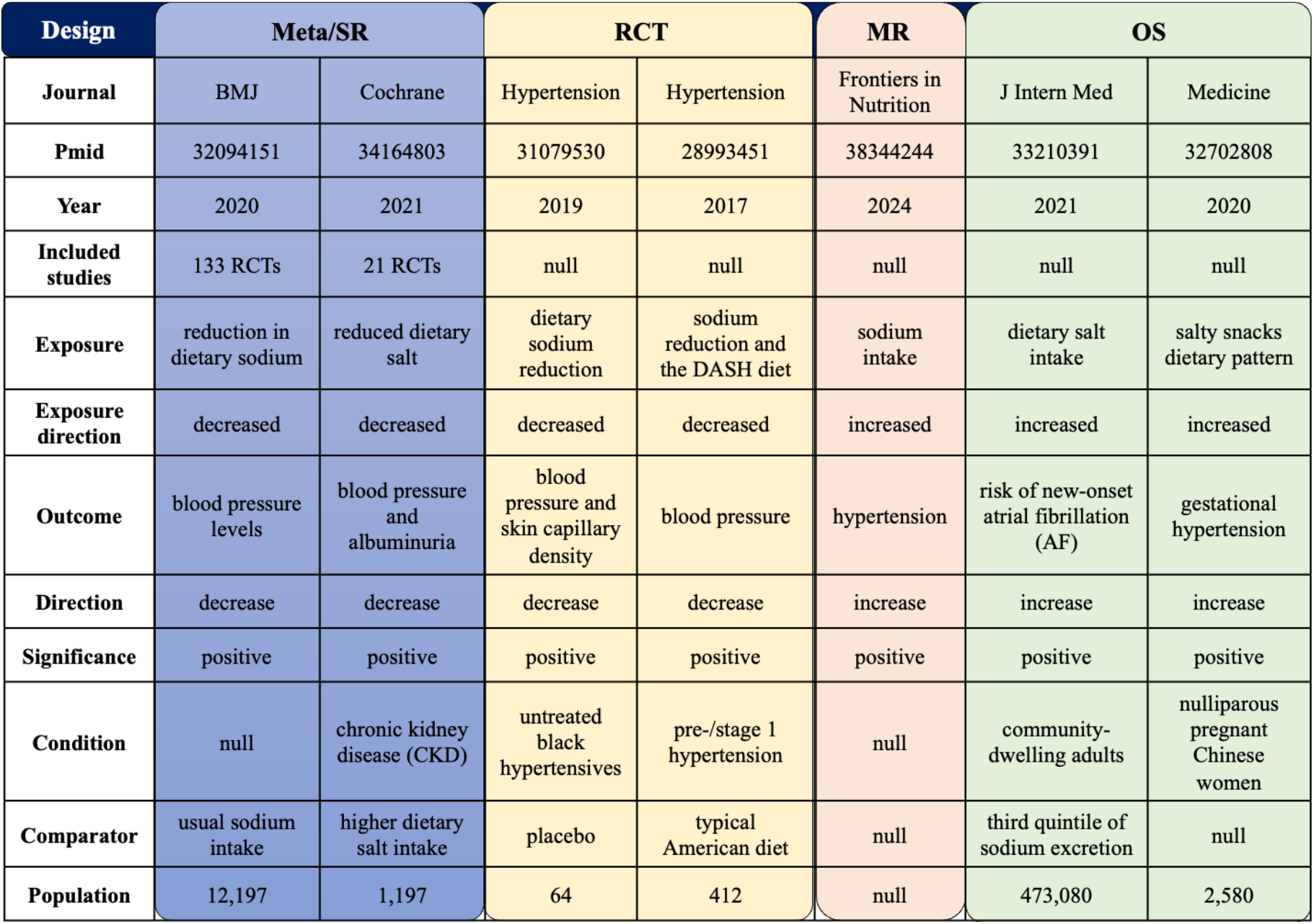
Example of automatic-extracted ready-for-triangulation evidence dataset of salt-on-hypertension.

To quantitatively triangulating the extracted evidence, we firstly designed an additional prompt asking LLM to identify if the extracted exposure/intervention and outcome match the target pair, i.e., salt intake on blood pressure (Supplementary file #4). We then removed irrelevant and only kept the matched extracted results. In this procedure, we also only included primary study designs (RCT, MR, and OS). Eventually 882 primary studies (325 RCTs, 3 MRs, and 554 OSs) with 1,931 extracted results are included in the following analysis.

In assessing the effect of salt intake on blood pressure, the LoE score indicates a moderate association with CoE 0.313 between increased salt intake and higher blood pressure (Figure 3). Interestingly, most OSs report increased salt intake, while RCTs primarily involve decreased salt intake. This difference reflects the varying definitions of intervention and exposure across study types and emphasizes the importance of triangulating evidence to minimize bias from the directionality of the factor under study.

**Figure 3.**
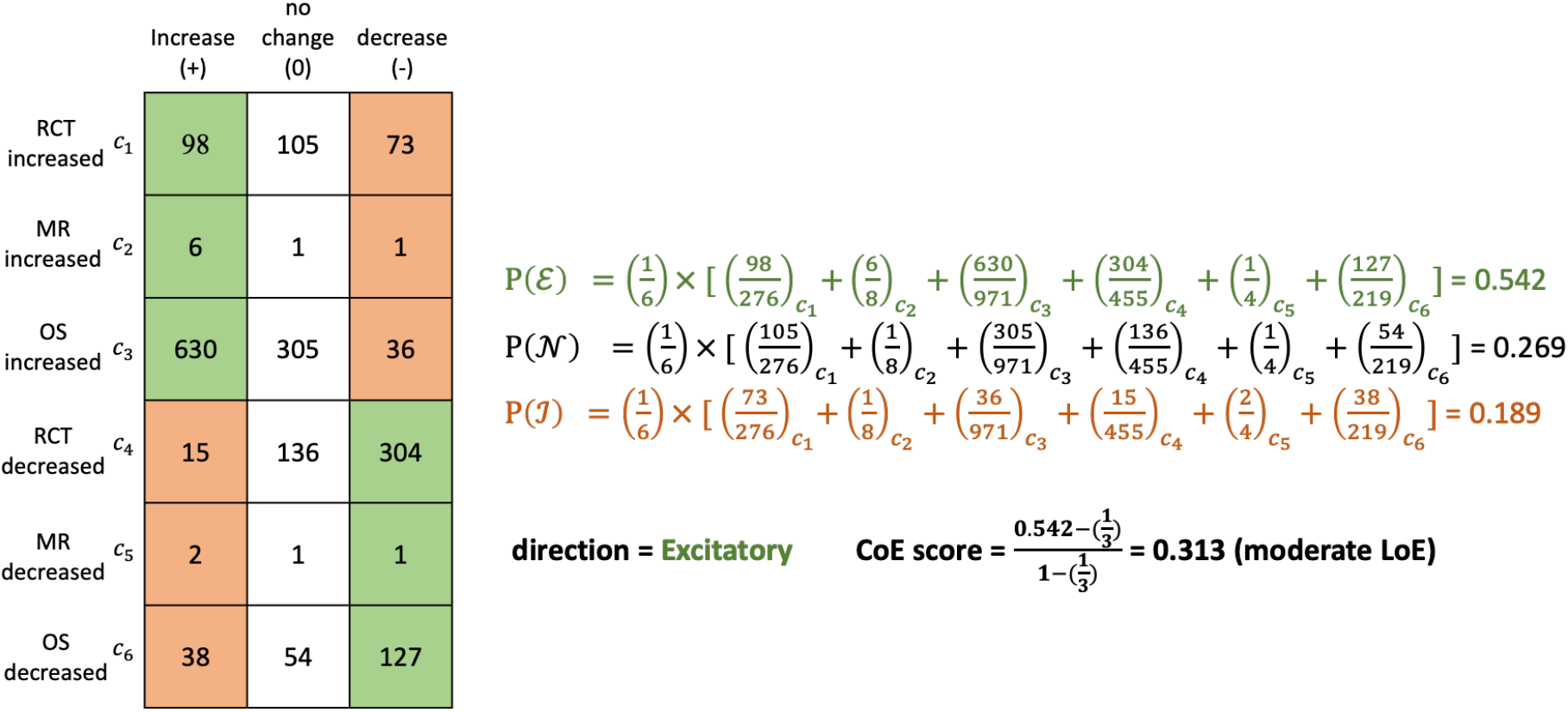
Convergency and Level of Evidence calculation for the case of salt intake on blood pressure. A convergency score of 0.313 reflects the moderate evidence supporting the excitatory relationship between salt intake and blood pressure. Green area and ℰ denote excitatory, white area and 𝒩 denote no change, green area and ℐ denote inhibitory.

We lastly compared the triangulation result with LoE with results extracted from meta-analyses, systematic reviews and reviews that were not included (Figure 4). In general, both results from triangulation and existing reviewing studies are consistent on the excitatory relationship between salt intake and blood pressure. A majority of existing studies focused on the relationship between decreased salt intake and blood pressure (151/176), and they concluded with an excitatory result in general, which is consistent with triangulated conclusion in this study. Nevertheless, there is a lack of reviewing studies focusing on increasing salt intake on blood pressure, possibly from primary OS and MR studies. Overall, our evidence triangulation method is consistent with existing meta-analyses and systematic review results, while also addressing the limitation of meta-analyses being constrained to a single type of study design.

**Figure 4.**
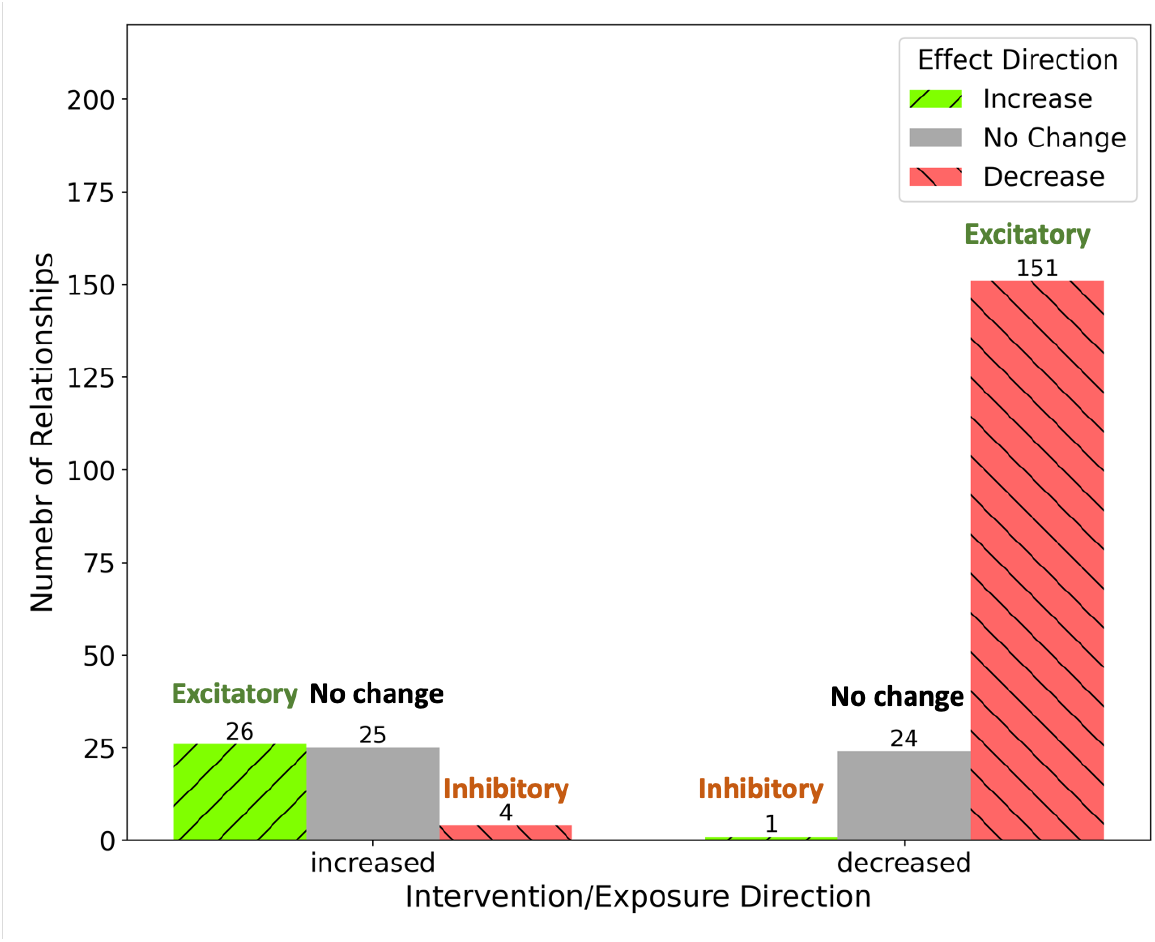
Number of relationships in meta-analyses, systematic reviews, and reviews of salt intake on blood pressure.

## Discussion

This study illustrates the potential of using LLMs to automate the extraction and triangulation of SDoH-related evidence across diverse study designs. Our approach utilizes a two-step extraction method, which first sequences exposure-outcome concept extraction followed by relation extraction, exhibiting better performance over the one-step method. This strategy essentially functions as a triple extractor, capturing entities and the relationships between them, similar to the SemRep system that extracts Subject-PREDICATE-Object semantic triples from biomedical text using a rule-based approach. The related ASQ platform has provided a user-friendly way to query SemRep-extracted triples along with associated evidence sentences, contributing to a novel approach for evidence triangulation. However, the extraction performance of SemRep is limited due to its reliance on a rule-based method developed two decades ago, despite ongoing updates and extensions with relation classification approaches^20-22^. Through data evaluations using CoE and LoE with a focused case study on the impact of salt intake on blood pressure, we demonstrated that LLMs can significantly simplify the synthesis of medical evidence, enhancing the efficiency of evidence-based decision-making. After triangulating evidence from different study designs, the relationship between salt intake levels and the risk of hypertension tends to indicate an excitatory effect direction. However, the level of evidence for this effect is low, suggesting that there are still many contradictory research findings. This result is consistent with the evidence status of salt controversy discussed in recent years^23, 24^.

This study also distinguishes our approach from others that utilize LLMs for meta-analysis. While decision-making should ideally be grounded in causal relationships between interventions and outcomes, predictions can be based on correlative relationships. Our objective was not to automate meta-analysis, which focuses on evidence derived from the same study design, such as RCTs. Recent proof-of-concept studies have shown that LLMs like Claude 2, Bing AI, and GPT-4 can improve the efficiency and accuracy of data extraction for evidence syntheses ^25-27^. However, these studies often involve limited datasets, either based on a single case^27^ or a small sample size of RCTs ^25, 26^. Although key data elements necessary for evidence triangulation, such as primary outcomes and effect estimates, can be accurately extracted in over 80% of RCTs, the performance of LLMs remains suboptimal with larger datasets. Additionally, other studies have evaluated the sensitivity and specificity of LLMs like GPT-3.5 Turbo in tasks such as title and abstract screening. The findings suggest that while these models offer promise, they are not yet sufficient to replace manual screening entirely^28^. On the other hand, GPT-4 Turbo-assisted citation screening has shown potential as a reliable and time-efficient alternative to systematic review processes ^29^. With these technological advances, Mayo Clinic has proposed an AI-empowered integrated framework for living, interactive systematic reviews and meta-analyses, enabling continuous, real-time evidence updates^30, 31^.

In contrast, our focus is on utilizing LLMs to perform convergency analysis among results obtained from different study designs, known as triangulation analysis. The key difference between meta-analysis and triangulation analysis lies in their focus: while meta-analysis assesses consistency within a single study design, triangulation analysis examines the convergency of conclusions across diverse study designs. Although there are no widely accepted quantitative methods for assessing convergency, insights can be drawn from convergency analysis, originated from neurobiological studies, primarily using a vote-counting approach across different study designs ^32-34^. Lastly, in causal graphical models, the concept of a causal relationship is uniform across different types of variables. Whether the graph pertains to biological or economic phenomena, the underlying principles of causality remain the same ^35^. This perspective also applies to the method in this study, that evidence should be and could be triangulated with CoE and LoE to conclude a causal relationship, whether in biomedical, economic, and environment field. As a result of constantly evolving research, the CoE ratings may change as more research findings becomes available. This is particularly the case for exposure-outcome pairings with low LoE due to contradictory results. Our approach is to harmonize confusion and help consumers make informed decisions about diet, exercise, and other activities that can affect their long-term health, as well as help researchers shape future clinical studies.

### Limitations

This study faced several limitations, including difficulties in accurately classifying study designs and interpreting associations due to data inconsistencies. The extracted entities were not mapped to standard biomedical vocabularies like SNOMED CT^36^ or UMLS^37^, leading to potential misalignment and incorrect relationship pairing, which could affect the final LoE. Furthermore, not all relevant study designs were included, limiting the comprehensiveness of the conclusions. The reliance on expert annotations also introduced subjective bias, potentially affecting the generalizability of the findings. While the two-step extraction approach showed improved performance, it requires further refinement to handle the complexity and variability of biomedical data effectively.

To maximize the utility of LLMs in evidence triangulation, future work should focus on addressing these limitations through continuous model fine-tuning and the development of more objective evaluation methods. A critical challenge remains in harmonizing evidence, including standardizing study populations and cause-and-effect entities across different study designs. The study aims to use LLMs to extract and align these elements through concept similarity measures like BERTScore, rather than relying on superficial string-based matches. Future studies will also introduce biomedical ontologies to better map the hierarchical structure of cause-and-effect concepts, leading to a more standardized and comprehensive approach to evidence triangulation.

## Methods

Our procedure begins by collecting titles and abstracts from relevant literature. We then apply a LLM to systematically process these texts across various study designs, extracting key outcomes and methodological details. This leads to the aggregation of data into a coherent, transparent dataset that is ready for triangulation analysis. The workflow ends with a quantitative evidence triangulation algorithm to discover the level of evidence behind a relationship between a SDoH factor and a health outcome.

### (1) Data sources

#### Validation dataset #1

Regarding data sources, the study utilizes literature categorized under publication types marked as meta-analysis, systematic reviews, observational studies, randomized controlled trials, clinical trials and related types available on PubMed. The MeSH terms “cardiovascular diseases” and “Diet, Food, and Nutrition” are utilized as search terms, with MeSH major topic as the search field. The resulting search query is outlined below: “*(cardiovascular diseases[MeSH Major Topic]) AND (Diet Food,and Nutrition[MeSH Major Topic])*”. For studies employing mendelian randomization (not a conventional publication type in PubMed), we additionally narrowed down the search to include only publication titles and abstracts containing the phrase “Mendelian randomization”. In total, 4,268 articles were retrieved. This first dataset will consist of 100 randomly selected studies from the corpus, used to validate the results extracted by LLM. The extracted results dataset and validation dataset are provided in supplementary data 1&2. Entities and relationships are manually annotated by 4 domain experts and clinicians (see Acknowledgement for details).

#### Validation dataset #2

The dataset used for external validation consists of 291 human-extracted relationships between dietary factors and coronary heart disease, derived from the Nurses’ Health Study^38^. It includes a wide range of dietary exposures, such as specific nutrients and food items, and their associations with cardiovascular outcomes. This data was meticulously curated and visualized in a knowledge graph, capturing both positive and negative associations, as well as effect size (hazard ratio, risk ratio, odds ratio, etc.) which can be served as a critical external foundation for testing the two-step extraction approach. The extracted results dataset and validation dataset are provided in supplementary data 3&4.

#### A Pilot-study dataset

To provide a specific example of the relationship between a particular disease and dietary factors, we further selected salt intake and hypertension as the intervention-outcome pair and retrieved relevant publications. Consistent with the aforementioned limitations on publication types, we refined the search terms to include MeSH terms related to salt intake and hypertension. The full constructed search queries are provided in Supplementary Material #2. After removing duplicates, we retrieved a total of 1,488 primary research articles. This case study dataset will be used to exhibit the formation of automatic-extracted ready-for-triangulation evidence dataset in the results section. The extracted results data is provided in supplementary data 5.

### (2) LLM-based study results extraction

For the task of extracting precise and insightful results from health-related documents, we employed medium-tier open source LLMs, which includes **deepseek-chat**^**39**^, **glm-4-airx**^**40**^, **qwen-plus**^**41**^, and **GPT-4-mini**^**42**^. While these models were not the top performers in all metrics, they were selected as a compromise, balancing both extraction performance and economic accessibility, such as cost per token. This balance makes them suitable for large-scale extraction tasks where both accuracy and cost-effectiveness are critical. The pricing of each model is shown in Supplementary Table 1.

The specific extraction tasks for the model are designed as following:

#### Methodological information

- **Identification of study design** The initial step involves using LLM to categorize the study design present in medical abstracts. The designs considered include RCT, MR, OS, and META.
- **Extraction for meta-analyses and systematic reviews** For abstracts identified as META, we ask LLM to extract the number of included studies and their respective study designs. This step is crucial for understanding the strength and diversity of evidence in these comprehensive analyses.

#### Ontological information

- **Primary result identification** Next, we ask LLM to identify the primary result from each abstract. This involves recognizing the main findings that the study reports, which is essential for summarizing the study’s major contribution to the field.
- **Intervention/Exposure and outcome extraction** Following the identification of the primary results, the model extracts key entities including intervention or exposure and the corresponding primary outcome. The model also identifies the direction (increased or decreased) of intervention/exposure for later relationship alignment.
- **Relationship and statistical significance** First the model extracts the direction of the relationship from the intervention/exposure to the outcome. The model assesses whether the intervention/exposure increases, decreases or an effect was not found. Then we ask LLM to extract statistical significance of the identified relationship, ensuring the ability to distinguishing positive results from negative results.
- **Population, Participant Number and Comparator Group information** Adhering to the standard representation medical evidence, we ask the model to extract information on the population condition under study, the number of participants, and details of the comparator group if applicable.

This prompt is to follow a logical progression from study-level information (study design), to more specific study result extraction (intervention/exposure, primary outcome, relationship direction, statistical significance), then contextual details (population, participant number, comparator). Figure 5 shows a graphical illustration of the overflow and logics of the designed prompt. For each abstract, LLM first determines the study design. If the abstract pertains to a meta-analysis, the model then identifies the number and types of included studies. Subsequently, it locates the primary result, extracts relevant details about the intervention/exposure and outcome, and assesses the direction and significance of the relationship. Information about the study population, the number of participants, and comparator group details are also extracted, providing a comprehensive overview of each study’s evidence.

**Figure 5.**
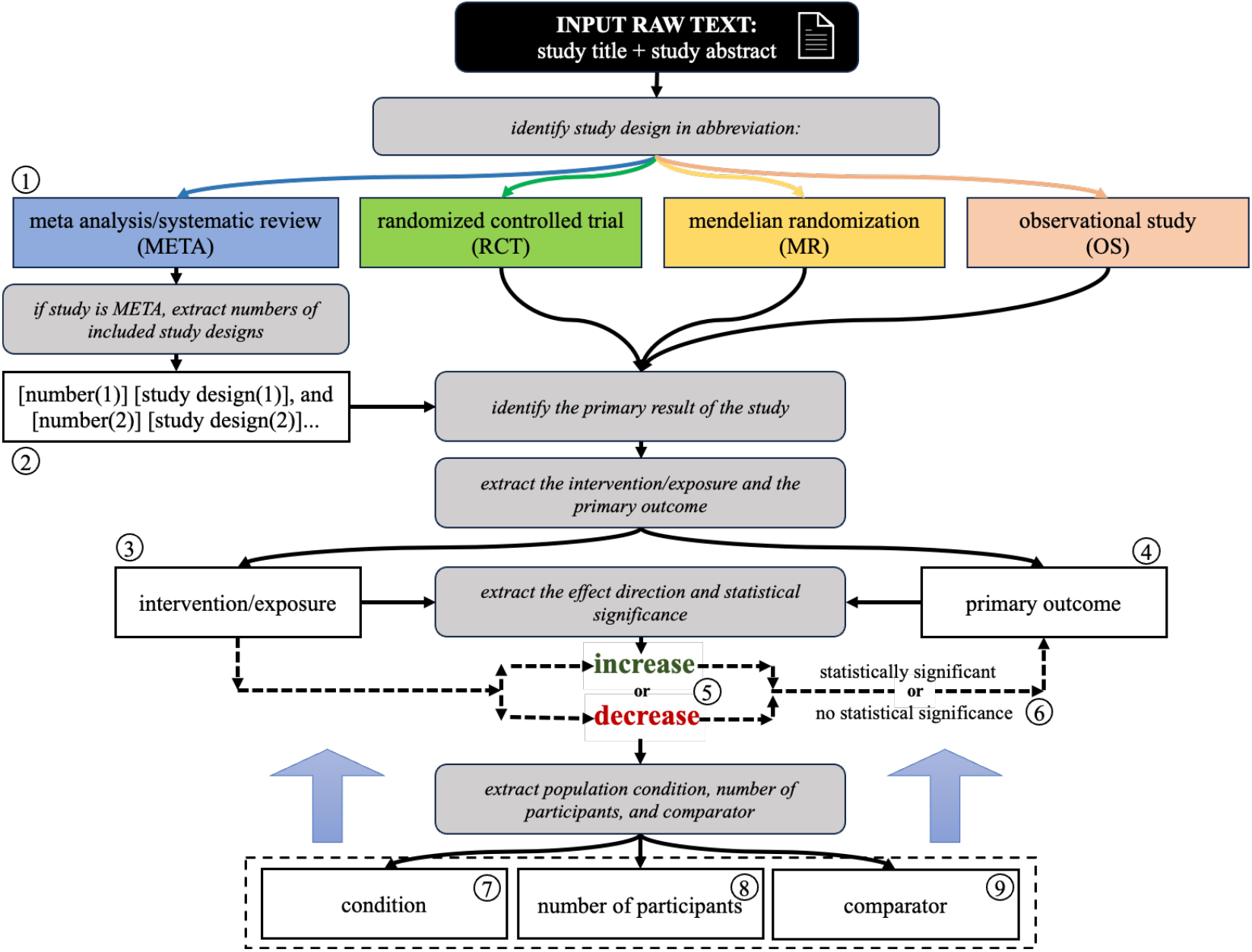
A flowchart describing the overall logic of using LLM to extract medical evidence in structured format. Gray boxes represent each part of the prompt in each step. Circled numbers (1-9) represent the extracted information by the model.

To enhance the accuracy and robustness of entity and relationship extraction, we implemented and compared a two-step extraction pipeline with a direct one-step extraction approach (Figure 6). The one-step extraction method simultaneously identifies and extracts both entities (e.g., exposures and outcomes) and their relationships directly from the text. In contrast, the two-step extraction process separates these tasks: the first step involves using NER to identify and extract entities from the text, such as specific dietary factors and cardiovascular outcomes. In the second step, these extracted entities are then used to identify and extract the relationships among them using RE techniques. This sequential approach allows for more precise entity recognition before relationship extraction, potentially reducing errors and improving overall extraction accuracy. Full prompts and code implementations for both the one-step and two-step extraction methods are detailed in the Supplementary Material #3, providing a comprehensive guide for replicating these processes.

**Figure 6.**
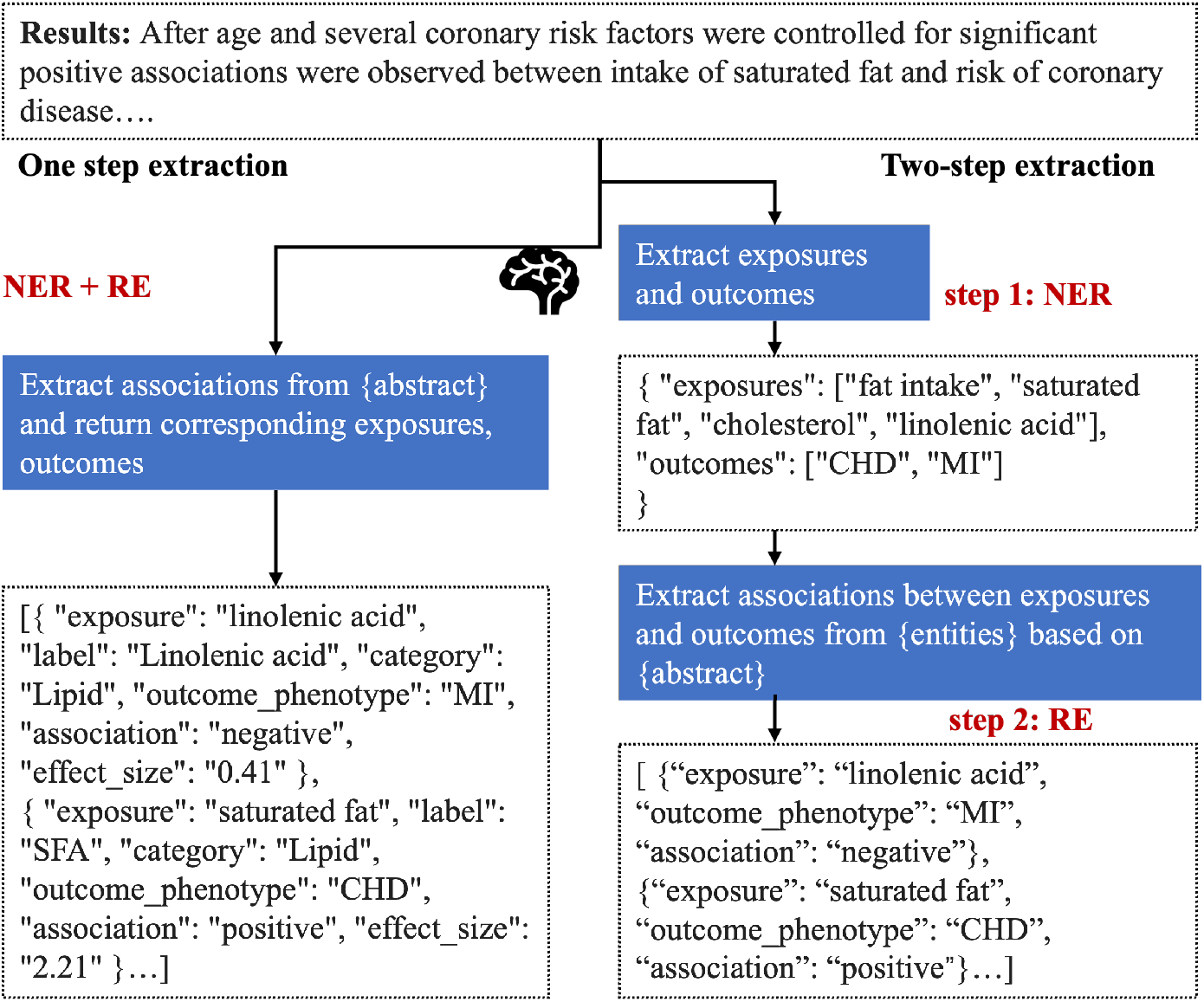
An illustration of one-step and two-step NER and RE in study results.

### (3) Convergency and level of evidence

To quantitatively analyze the dataset, we developed an evidence triangulation algorithm derived from the Cumulative Evidence Index (CEI) score from ResearchMap, which is a graph-based representation of empirical evidence and hypothetical assertions found in research articles, allowing biologists to systematically evaluate and plan experiments^32-34^. The scoring algorithm uses a Bayesian approach to evaluate the strength of evidence for a causal relationship in research maps by integrating results from various types of experiments, accounting for convergency and consistency. Originally, CEI calculates the amount of evidence of relationships between agents(source) and targets(outcome) across different study designs including positive interventions, positive non-interventions, negative non-interventions, and negative interventions.

In this study, we optimized the CEI algorithm for population-based health studies and proposed CoE. CoE score is calculated by first categorizing studies into different classes based on primary study designs and exposure/intervention directions (e.g., RCT with increased intervention or OS with decreased exposure). Then each study result is entered into a scoring table using Laplace smoothing, which adds a pseudo count to avoid zero denominators. The evidence is then tallied to determine how strongly each type of relationship (excitatory, inhibitory, or no change) is supported.

The final score is derived by normalizing the difference between the maximum observed evidence and a baseline prior, resulting in a value between 0 and 1 that reflects the strength and consistency of the evidence for a causal relationship. Each relationship is given a possibility score representing the average proportion across study designs, and the determinate relationship is the relationship with biggest possibility. The detailed Bayesian algorithm can refer to ResearchMaps^33^. In this study we calculated the CoE score for each relationship then categorized the score into 3 levels: weak [0-0.3], moderate (0.3-0.7), and strong [0.7-1.0], and named the scoring method as LoE to represent different levels of convergency in evidence.

## Data Availability

All data produced in the present study are available upon reasonable request to the authors

## Acknowledgement

This study was funded by the National Key R&D Program for Young Scientists (Project number 2022YFF0712000 to JD) and the National Natural Science Foundation of China (Project number 72074006; 82330107 to JD). We declare no conflicts of interest.

We are grateful to the following experts for their invaluable contributions and insightful feedback on this study: Dr. Guohua He from Sun Yat-sen University First Affiliated Hospital, Dr. Na He from Peking University Third Hospital, Dr. Zhenhua Lu from Peking University Cancer Hospital, Dr. Weihua Hu from Peking University, and Dr. Mingming Zhao from Peking University Third Hospital.

## Author Contributions

Conceptualization: J.D., X.S.

Methodology: J.D., X.S., C.Y., T.C.

Formal Analysis: X.S.

Data Curation: X.S., W.Z., T.C.

Writing—original draft: X.S., J.D.

Writing—review & editing: J.D.

Funding acquisition: J.D.

